# Real-time Estimation of Global CFR Ascribed to COVID-19 Confirmed Cases Applying Machine Learning Technique

**DOI:** 10.1101/2021.12.27.21268463

**Authors:** Monalisha Pattnaik, Aryan Pattnaik

## Abstract

The COVID-19 is declared as a public health emergency of global concern by World Health Organisation (WHO) affecting a total of 201 countries across the globe during the period December 2019 to January 2021. As of January 25, 2021, it has caused a pandemic outbreak with more than 99 million confirmed cases and more than 2 million deaths worldwide. The crisp of this paper is to estimate the global risk in terms of CFR of the COVID-19 pandemic for seventy deeply affected countries. An optimal regression tree algorithm under machine learning technique is applied which identified four significant features like diabetes prevalence, total number of deaths in thousands, total number of confirmed cases in thousands, and hospital beds per 1000 out of fifteen input features. This real-time estimation will provide deep insights into the early detection of CFR for the countries under study.

**CFR:** 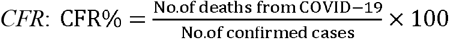as suggested by (Boldog et al., 2020, Chakraborty et al. 2019, Russell et al., 2020)

**Diabetes Prevalence:** proportion of a population who have diabetes in a given period of time.

**Stringency Index:** it provides a computable parameter to evaluate the effectiveness of the nationwide lock down in a particular country.

**GDP Per Capita:** it is a metric that breaks down a country’s economic output per person and is calculated by 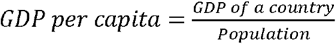

**Population Density:** it is a measurement of population per unit area. It refers to the number of people living in an area per square kilometre.

**HDI:** it is a statistic composite index of life expectancy, education (literacy rate, gross enrolment ratio at different levels and net attendance ratio) and per capita income indicators which are used to rank countries into four tiers of human development.

## 1. Introduction

The identification of the factors contributing to the global risk estimation due to the confirmed COVID-19 cases has become a challenging task for all the countries and territories in the world. During the present unending phase of global pandemic due to COVID-19, it is highly important to estimate CFR accurately. The estimates of CFR are highly dependent on country-specific demographic parameters, adequate health care facilities, different COVID-19 symptoms, epidemiological characteristic, diabetes and cardiovascular disease. The present study is undertaken to identify the most important factors causing the CFR of the selected countries using an optimal regression tree model (Breiman et al., 1984). The regression tree in machine learning follows a built-in mechanism with a set of numerical, categorical and integer input variables to model the arbitrary decision boundaries. The regression tree method finds an exceptional classification of variables than that of the classification models. This RT is a target specific machine learning technique which encompasses recurrent binary splitting of the variables to produce a tree structure which is followed by a trimming process of removing insignificant variables. Hence, finally finds the variables having significant contribution on CFR.

The rest of the paper is structured as stated below. Section 2 deals with country wise datasets on CFR and the independent variables and machine learning regression tree technique. Findings and discussions are presented in Section 3. Finally, Section 4 deals with conclusion and recommendations.

## 2. Global Risk Assessment of COVID-19 Confirmed Cases

Initially of the COVID-19 outbreak, country-wise data on case fatality rates due to COVID-19 and other features related to this were obtained for 70 profoundly affected countries. The case fatality rate of the country is plainly defined as the number of deaths due to COVID-19 divided by the confirmed number of COVID-19 cases of that country. This section explains about the list of essential input variables that have well-built influences on the CFR of the country.

### 2.1 Country wise Datasets

In the pace of rapidly changing data for COVID-19, we calculated the case fatality ratio estimates for 70 selected countries (see Figure 1) from the day of starting the outbreak to 25 January 2021 from the following website^1^. A lot of first round analysis is done to determine a set of input variables, some of which are expected to be critical causal variables for risk estimating of COVID-19 in these affected countries. Studies (Boldog et al., 2020, Jung et al., 2020, Nishiura et al. 2020, Gahan et al., Chakraborty et al. 2019, Russell et al., 2020) have suggested that the total number of cases, age distributions, and shutdown period have high impacts on the CFR values for some of the countries. Along with these three variables, we also considered twelve more demographic structures and disease characteristics for these countries as input variables that are likely to have a potential impact on the CFR estimates. Therefore, the CFR modeling dataset consists of 70 observations having fifteen possible causal or input variables and one numerical response or output variable (CFR), as reported in Table 1. The potential causal variables considered for global risk estimation are the followings: the total number of COVID-19 cases (in thousands) in the country till 25 January, 2021, total number of deaths (in thousands), stringency index, population (in millions), population density per *km*^2^ for the country, median age, aged 65 older, aged 70 older, GDP per capita, cardiovasc death rate, diabetes prevalence, hospital beds per 1000, life expectancy, HDI, time period (in days) count (from the starting day of COVID-19 cases for the country to January 25 2021) of the country. The dataset contains a total of 10 numerical input variables and two integer input variables.

**Table 1:** Descriptive statistics of control variables and response variable of CFR dataset of 70 countries

| Sl. No. | Input (Control) and Output (Response) Variables of 70 no. of Countries | Variable Type | Min | Max | Mean | First Quartile | Median | Third Quartile | Standard Deviation |
| --- | --- | --- | --- | --- | --- | --- | --- | --- | --- |
| 1 | Total cases in thousand | Integer | 54.672 | 25297.059 | 1366.536 | 196.642 | 395.424 | 1070.268 | 3386.693 |
| 2 | Total deaths in thousand | Integer | 2.011 | 421.129 | 29.853 | 3.400 | 8.946 | 24.658 | 61.41977 |
| 3 | Stringency index | Numerical | 30.56 | 89.81 | 68.37 | 61.69 | 70.37 | 76.39 | 11.71370 |
| 4 | Population (millions) | Integer | 2.079 | 1439.0 | 88.01 | 9.17 | 19.18 | 64.07 | 237.5685 |
| 5 | Population density | Numerical | 4.037 | 1256.036 | 136.149 | 54.600 | 90.615 | 137.012 | 174.3895 |
| 6 | Median age | Numerical | 18.60 | 48.20 | 35.02 | 29.10 | 35.50 | 41.95 | 7.976771 |
| 7 | Aged 65 older | Numerical | 2.581 | 27.049 | 11.952 | 6.066 | 10.976 | 18.558 | 6.444895 |
| 8 | Aged 70 older | Numerical | 1.337 | 18.493 | 7.738 | 3.839 | 6.947 | 11.952 | 4.536677 |
| 9 | GDP per capita | Numerical | 1730 | 67335 | 22270 | 9946 | 17950 | 32305 | 15584.57 |
| 10 | Cardiovasc death rate | Numerical | 79.37 | 597.03 | 245.01 | 134.98 | 206.28 | 321.76 | 132.5009 |
| 11 | Diabetes prevalence% | Numerical | 3.280 | 17.720 | 7.627 | 5.835 | 7.155 | 8.505 | 2.646095 |
| 12 | Hospital beds per 1000 | Integer | 0.300 | 13.050 | 3.450 | 1.425 | 2.650 | 5.405 | 2.57319 |
| 13 | Life expectancy | Numerical | 64.13 | 84.63 | 76.44 | 73.64 | 76.68 | 80.75 | 4.821063 |
| 14 | HDI | Numerical | 0.4630 | 0.9440 | 0.784 | 0.7103 | 0.7900 | 0.8775 | 0.1125366 |
| 15 | Time period (in days) | Integer | 305 | 391 | 342.1 | 329 | 335.5 | 360 | 18.41732 |
| 16 | CFR %(Response Variable) | Numerical | 0.7334 | 8.4817 | 2.4299 | 1.5901 | 2.122 | 2.7883 | 1.328249 |

**Figure 1:**
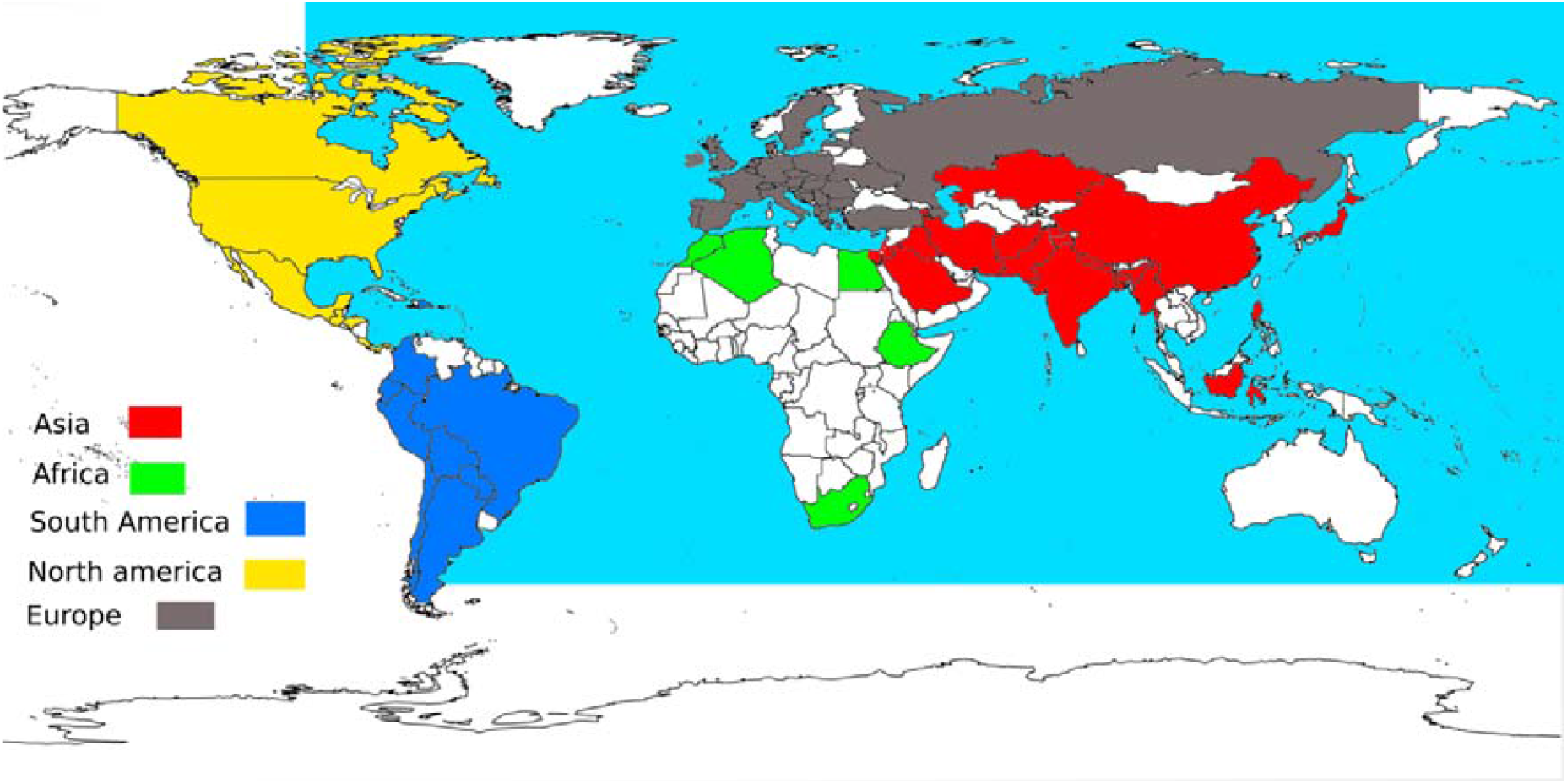
Seventy deeply COVID-19 affected countries

### 2.2 Regression Tree Method

For the estimation of global CFR dataset for 70 countries, the regression tree (RT) (Breiman et al., 1984) is applied that has built-in feature selection mechanism, easy interpretability, and provides better visualization. RT, as a widely used simple machine learning algorithm, can model arbitrary decision boundaries. The methodology outlined in (Breiman et al., 1984) can be summarized into three stages. The first stage involves growing the tree using a recursive partitioning technique to select essential variables from a set of possible causal variables and split points using a splitting criterion. The standard splitting criteria for RT is the mean squared error (MSE). After a large tree is identified, the second stage of RT methodology uses a pruning procedure that gives a nested subset of trees starting from the largest tree grown and continuing the process until only one node of the tree remains. The cross-validation technique is popularly used to provide estimates of future prediction errors for each subtree. The last stage of the RT methodology selects the optimal tree that corresponds to a tree yielding the lowest cross-validated or testing set error rate. To avoid instability of trees in this stage, trees with smaller sizes, but comparable in terms of accuracy, are chosen as an alternative. This process can be tuned to obtain trees of varying sizes and complexity. A measure of variable importance can be achieved by observing the drop in the error rate when another variable is used instead of the primary split. In general, the more frequent a variable appears as a primary split, the higher the importance score assigned. A detailed description of the tree building process is available at (James et al., 2013).

## 3. Findings and Discussions

The motivation behind the choice of RT as a potential model to find the significant input variables out of 15 input variables for the CFR estimates of the country is the simplicity, easy interpretability, and high accuracy of the RT algorithm. We apply an optimal RT model to the dataset consisting of 70 different country samples and try to find out potential casual variables from the set of available variables that are related to the case-fatality rates of the country. RT is implemented using ‘rpart’ (Therneau et al., 2015) package in R with “minsplit” equals to 10% of the data as a control parameter. We have used RMSE, MAPE, coefficient of multiple determination (R^2^), and adjusted R^2^ (AdjR^2^) to evaluate the predictive performance of the tree model used in this study (James et al., 2013). An optimal regression tree is built with 4 variables with ‘minsplit’ = 5 with equal costs for each variable. The estimates of the performance metrics for the fitted tree are as follows: RMSE = 0.7151, MAPE=0.1827, R^2^ 0.7059, and R^2^ 0.6878. A variable importance list from the RT is given in Figure 2 and the fitted tree is provided in Figure 3. From the variable importance plot based on the complexity parameter of the RT model (also see Figure 3), four causal variables are obtained out of 15 potential input variables having higher importance. Our results are consistent with previous results obtained by (Boldog et al., 2020, Jung et al., 2020, Chakraborty et al. 2020, Russell et al., 2020), where the authors suggested that the total number of cases, age distributions, people of age group > 65 years, hospital beds per 1000 people and shutdown period have high impacts on the CFR estimates. But interestingly, we obtained four essential causal variables like diabetes prevalence, total number of deaths in thousands, total number of confirmed cases in thousands and hospital beds per 1000 people can be managed to fight against this deadly disease. Once these variables are taken care of, the respective country may reduce their case fatality rate at a significant rate.

**Figure 2:**
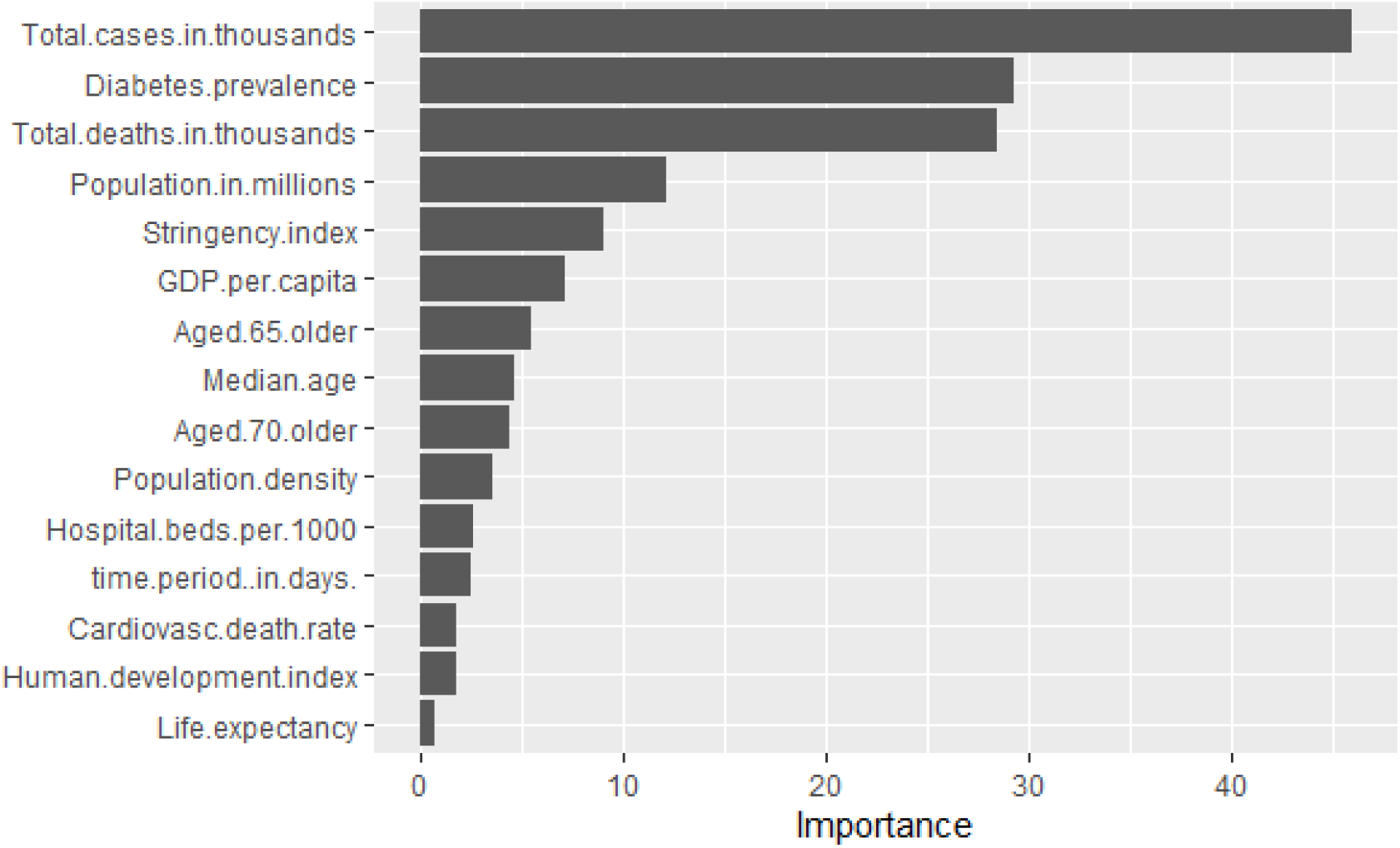
Feature importance percentages affecting the CFR based on a complexity parameter applying Regression Tree (RT)

**Figure 3:**
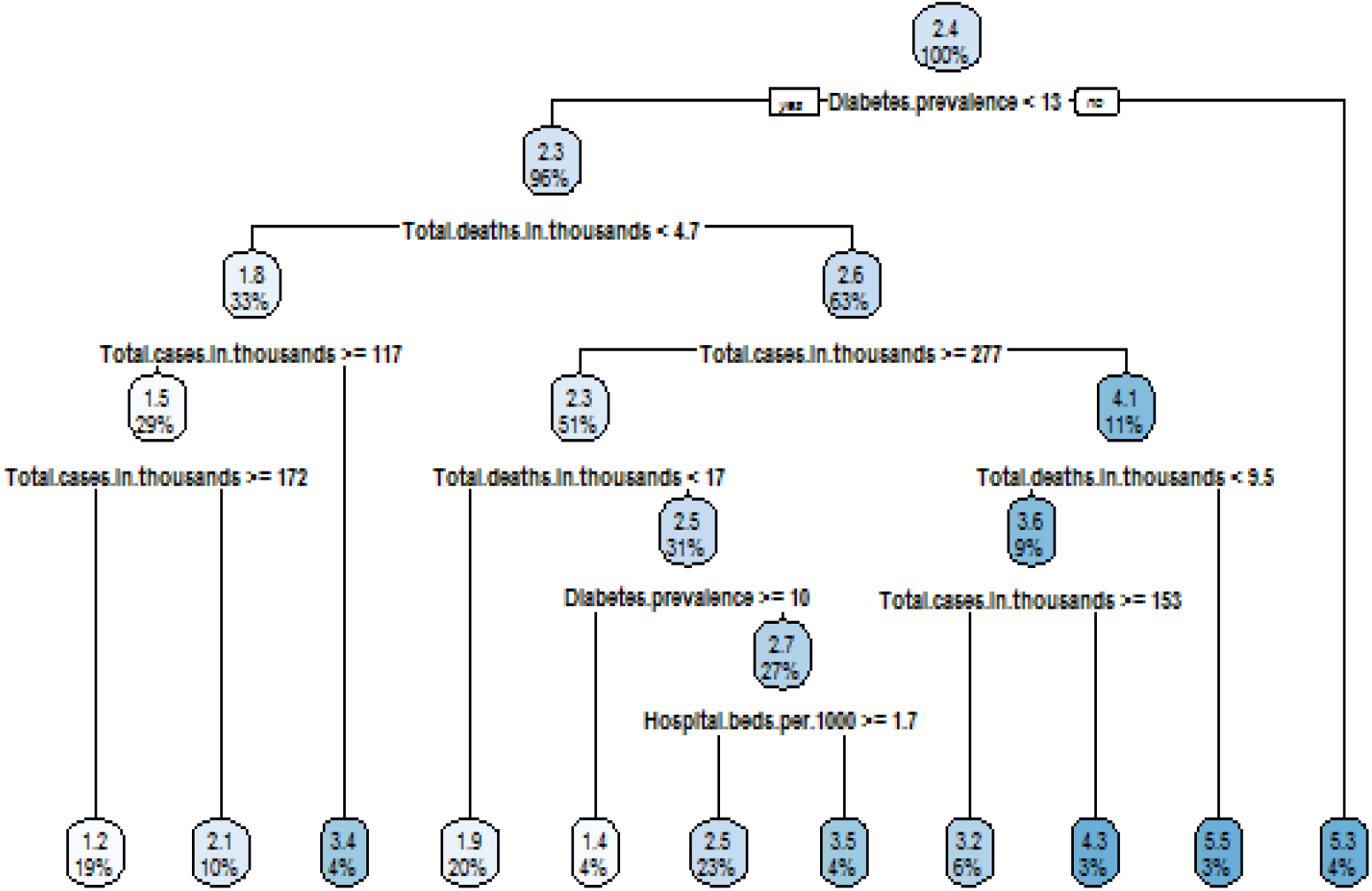
Optimal tree representing the relationships between the causal variables and CFR

By applying rpart the range of cost complexity can be evaluated. To compare the error for each cost complexity value rpart performs a 10-fold cross validation so that the error associated with a given cost complexity is computed on the hold-out validation data. Figure 3 shows the optimal tree having 10 internal nodes resulting in 11 terminal nodes. Basically, this tree is partitioning on 4 variables to produce its model. A tree with 11 terminal nodes we can force to generate a full tree by using cp=0 (See Figure 4). In Figure 4 y-axis is cross validation error, lower x-axis is cost complexity value, upper x-axis is the number of terminal nodes. After 11 terminal nodes, we see diminishing returns in Error reduction as the tree grows deeper. To predict the CFR of a country based on diabetes prevalence, total deaths, total cases and hospital beds per 1000. All 70 number of countries go through the RT (see Figure 3), are assessed at a particular node, and proceed to the left if the answer is “yes” or proceed to the right if the answer is “no”. So, first all 70 countries that have less than 13% of diabetes prevalence go to the left branch, all other countries proceed to the right branch. All countries that greater than 13% of diabetes prevalence (far right branch) have average CFR is 5.3%. This branch leads to terminal nodes or leaf which contains the predicted response value of CFR. Hence, all the countries that have less than 13% diabetes prevalence, have less than 4.7 number of total deaths in thousands and have greater than 117 of total confirmed cases in thousands have average CFR is 1.2%. Whereas, all the countries that have less than 13% diabetes prevalence, have less than 4.7 number of total deaths in thousands and have lying between 117 and 172 number of total confirmed cases in thousands have average CFR is 2.1%. All the countries that have less than 13% diabetes prevalence, have less than 4.7 number of total deaths in thousands and have less than 117 number of total confirmed cases in thousands have average CFR is 3.4%. All the countries that have less than 13% diabetes prevalence, have lying between 4.7 and 17 number of total deaths in thousands and have greater than 277 number of total confirmed cases in thousands have average CFR is 1.9%. All the countries that have lying between 10% to 13% diabetes prevalence, have greater than 4.7 number of total deaths in thousands and have greater than 277 number of total confirmed cases in thousands have average CFR is 1.4%. All the countries that have less than 13% diabetes prevalence, have greater than 4.7 number of total deaths in thousands, have greater than 277 number of total confirmed cases in thousands and have greater than 1.7% hospital bed per 1000 have average CFR is 2.3% whereas have less than that of 1.7% hospital bed per 1000 have average CFR is 3.5%. All the countries that have less than 13% diabetes prevalence, have lying between 4.7 and 5.5 number of total deaths in thousands and have lying between 153 and 277 number of total confirmed cases in thousands have average CFR is 3.2% whereas have less than that of 277 number of total confirmed cases in thousands have average CFR is 4.3%. All the countries that have less than 13% diabetes prevalence, have greater than 4.5 number of total deaths in thousands and have greater than 277 number of total confirmed cases in thousands have average CFR is 5.5%. The splitting process continues, visiting all variables each time a split is made until all the CFR data are divided into eleven partitions with predicted CFRs (1.2, 2.1, 3.4, 1.9, 1.4, 2.5, 3.5, 3.2, 4.3, 5.5, 5.3) based on only four input variables namely diabetes prevalence, total number of deaths in thousands, total number of confirmed cases in thousands, and hospital beds per 1000 people.

**Figure 4:**
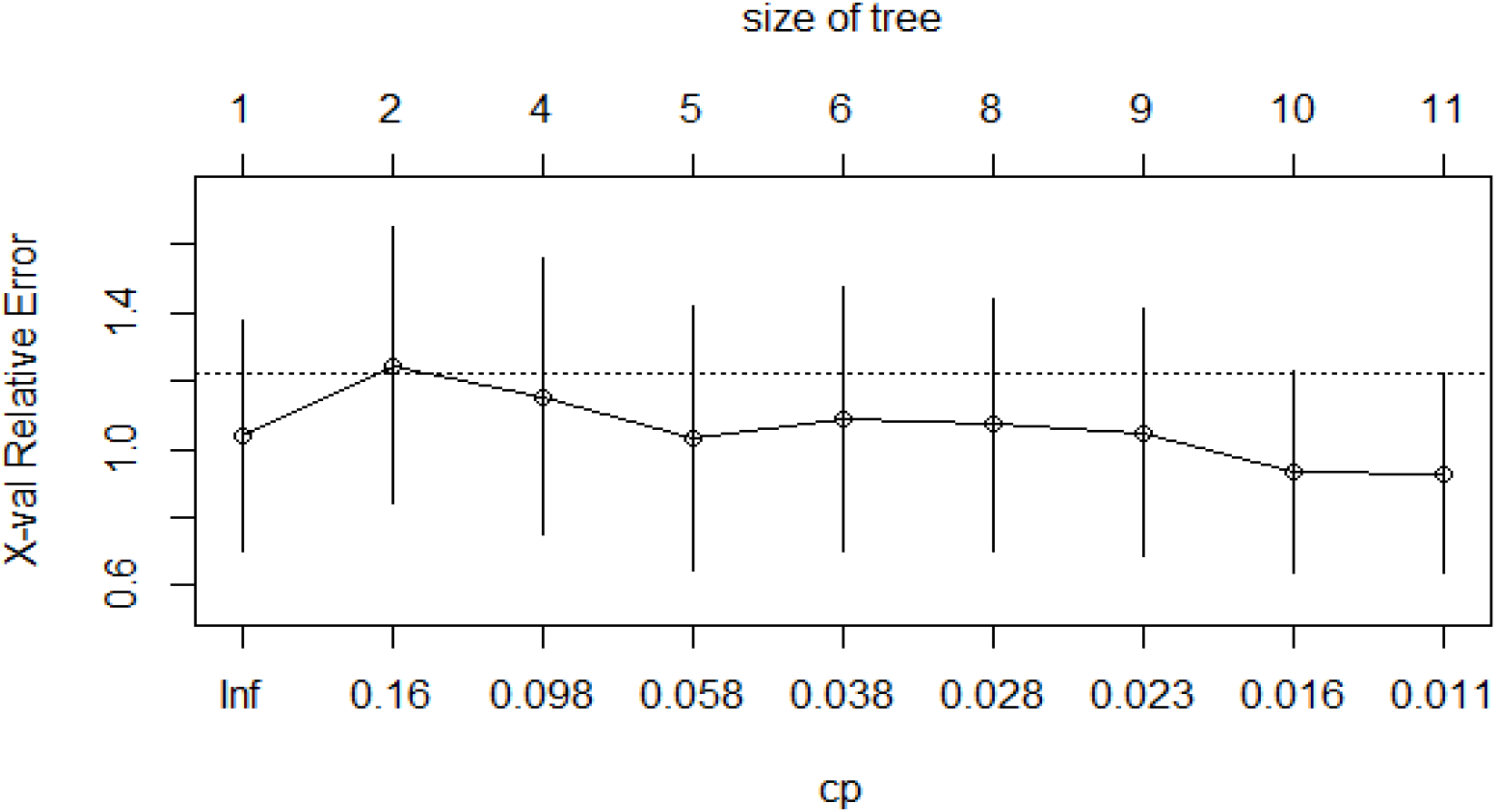
Complexity parameter and size of the tree verses cross validation error

The risk of COVID-19 is estimated by finding four key parameters that are expected to have strong associations with that of case fatality rates of the country. Applying machine learning approach, an optimal regression tree model is used to measure global risk for COVID-19 pandemic. The model is very flexible, easily interpretable, and the more data will come, one can just incorporate the new data sets and rebuild the trees to get the updated estimates. RT provides a better visual representation and is easily interpretable to be understood by a broader audience. Quantification of the outbreak risks and their dependencies on the key parameters will support the governments and policymakers for the planning of health care systems in different countries that faced this unending epidemic. Experimental results suggest four control variables out of fifteen highly influential variables that will have a significant impact on controlling CFR of the country. Below we present a point by point discussion of the control variables affecting CFR and preventive actions to be taken by the governments.

1. Controlling diabetes prevalence of the country can reduce CFR and number of deaths due to COVID-19.
2. Number of confirmed cases should be specially taken care of by using face mask, social distancing, hand washing, using sanitizer etc.
3. The number of hospital beds should be increased by making special health care arrangements in other places to deal with this emergency due to COVID-19.

## 4. Conclusions

We made some simplifying assumptions to carry out the analysis of COVID-19 datasets. The assumptions are listed as follows: (a) the virus mutation rates are comparable for different countries; (b) the recovered persons will achieve permanent immunity against COVID-19; (c) we ignore the effect of climate change. Along in this line, we presented approach to deal with one inter-connected problems on COVID-19. In this problem of risk estimation, we found some important factors affecting case fatality rates of COVID for 70 highly affected nations in the globe. However, there may exist a few more controllable factor(s), and some disease-based characteristics that can also have an impact on the value of CFR for different countries, can be regarded as future scope of the study.

## Data Availability

All data produced in the present study are available upon reasonable request to the authors

## Abbreviations and Meaning

COVID-19: Coronavirus Disease 2019
CFR: Case Fertility Rate
RT: Regression Tree
GDP: Gross Domestic Product
HDI: Human Development Index
CP: Complexity Parameter.

## Ethical approval

This study was based on publicly available data and did not require ethical approval.

## Footnotes

1 https://www.worldometers.info/coronavirus/

